# The Utility of rRT-PCR in Diagnosis and Assessment of Case-fatality rates of COVID-19 In the Iranian Population – Positive Test Results are a Marker for Illness Severity

**DOI:** 10.1101/2020.04.29.20085233

**Authors:** Ghasem Janbabaei, Eric J. Brandt, Reza Golpira, Alireza Raeisi, Jafar Sadegh Tabrizi, Hamid Reza Safikhani, Mohammad Taghi Talebian, Siamak Mirab Samiee, Alireza Biglar, Reza Malekzadeh, Arya Mani

## Abstract

The utility of PCR-based testing in characterizing patients with COVID-19 and the severity of their disease remains unknown. We performed an observational study among patients presenting to hospitals in Iran who were tested for 2019-nCoV viral RNA by rRT-PCR between the fourth week of February 2020 to the fourth week of March 2020. Frequency of symptoms, comorbidities, intubation, and mortality rates were compared between COVID-19 positive vs. negative patients. 96103 patients were tested from 879 hospitals. 18754 (19.5%) tested positive for COVID-19. Positive testing was more frequent in those 50 years or older. The prevalence of cough (54.5% vs. 49.7%), fever (49.5% vs. 44.7%), and respiratory distress (43.0% vs. 39.0%) but not hypoxia (46.9% vs. 56.7%) was higher in COVID-19 positive vs. negative patients (p<0.001 for all). More patients had cardiovascular diseases (10.6% vs. 9.5%, p<0.001) and type 2 diabetes mellitus (10.8% vs. 8.7%, p<0.001) among COVID-19 positive vs. negative patients. There were fewer patients with cancer (1.1%, vs. 1.4%, p<0.001), asthma (1.9% vs. 2.5%, p<0.001), or pregnant (0.4% vs. 0.6%, =0.001) in COVID-19 positive vs. negative groups. COVID-19 positive vs. negative patients required more intubation (7.7% vs. 5.2%, p<0.001) and had higher mortality (14.6% vs. 6.3%, p<0.001). Odds ratios for death of positive vs negative patients range from 2.01 to 3.10 across all age groups. In conclusion, COVID-19 test-positive vs. test-negative patients had more severe symptoms and comorbidities, required higher intubation, and had higher mortality. rRT-PCR positive result provided diagnosis and a marker of disease severity in Iranians.

## INTRODUCTION

Following the spread of the 2019 novel coronavirus (SARS-COV-2) in Asia, the first documented case of COVID-19 in Iran was made on February 12, 2020 in the City of Qom. The Deputy for Health and Curative Affairs at the Iranian Ministry of Health and Medical Education (MOHME) responded urgently by compiling World Health Organization (WHO) guidelines for COVID-19 prevention, diagnosis and treatment. The MOHME designed and launched its National Mobilization Plan Against COVID-19 in a collaborative effort between the Ministry’s Deputy for Health and Curative Affairs, military organizations, non-governmental organizations (NGOs), and other volunteers.

Relying on a centralized health care system, the health-care authorities of Iran initiated a coordinated effort to implement preventive measures and to diagnose and treat patients with COVID-19. Close to 1,000 comprehensive health centers functioning 16 and 24 hours per day, seven days of the week were designated to provide specific COVID-19 services according to a standard protocol. Immediate in-person education was provided to first line responders throughout the country in preparation for the spread of the coronavirus. Instruction for screening, diagnosis and treatment of COVID-19 was provided through widely available electronic National Health care system. An interactive website linked to medical records was designed and made widely accessible to the public for self-screening of the general population. The website can be accessed using national identification number and birth date and provides medical advice according to individual’s responses to specific disease-related questions.

In this process, it soon became apparent that without an efficient tool for the diagnosis there is a little to no chance to harness this fatal pandemic. Real-time reverse transcriptase–polymerase chain reaction (rRT-PCR) of nasopharyngeal swabs is widely used to confirm the clinical diagnosis, but significant false negative results have created concerns about its utility[1]. Additionally, it is unclear what presenting symptoms and comorbidities are more common amongst COVID-19 positive vs. negative patients.

Therefore, we completed a study to compare the frequency of presenting symptoms, comorbidities, mechanical ventilation, or death between COVID-19 positive and negative patients hospitalized with acute respiratory illness.

## METHODS

### Study population

We performed an observational study using data collected from patient registry records of individual hospitals in 879 designated hospitals from all 31 provinces of Iran between the fourth week of February to the fourth week of March.

### Case identification

As of the fourth week of March 2020, millions of individuals were screened either by phone or by self-evaluation using interactive websites. Screening including questions about specific symptoms, including fever, chills, cough, sore throat, respiratory distress, and their potential contact with individuals with suspected COVID-19 infection. All individuals were provided with information for prevention, modes of disease transmission and means of personal protection. Individuals identified as possible COVID-19 cases were referred to specialized health-care centers for COVID-19 testing. Simultaneously, these centers received a text message to contact the suspected patients for the initiation of diagnosis, home therapy, and subsequent follow-ups or referral to specialized centers. Those deemed as high risk for acquiring the disease, including those with T2D and hypertension, obesity, immunodeficiency, malignant disorders undergoing immune suppressive therapy, and pregnant women were prioritized. At the specialized health-care centers patients with severe respiratory distress and/or oxygen saturation below 93%, reduced levels of consciousness, or intractable cough were sent to COVID-19 referral hospitals for hospitalization. All hospitalized COVID-19 positive patients were put on a triple therapy treatment protocol that included Hydroxychloroquine (400 mg loading dose, followed by 200 mg twice a day), Lopinavir-Ritonavir (Kaletra), and Ribavirin for 2 weeks. Hemodynamically stable patients with mild symptoms were placed on Hydroxychloroquine and discharged to home quarantine. All data were entered into a computerized database at the National MOH COVID-19 Database.

### Assay

Diagnosis of COVID-19 was made based on the presence of viral RNA by real-time reverse-transcriptase–polymerase-chain-reaction (rRT-PCR) assays in accordance with the protocol established by the WHO[2] at the Pasteur Institute of Iran and at the National Reference Laboratories.

### Study design, definitions, and diagnosis

Data extracted includes clinical symptoms or signs, comorbidities, ventilator use, and mortality. Radiologic assessments included computed tomography (CT) of the chest, which were not available at the time of preparation of this manuscript.

Symptoms or signs included cough, fever, respiratory distress, hypoxia, myalgia, and reduced level of consciousness. Fever was defined as a forehead temperature ≥37.6°C. Hypoxia was defined as oxygen saturation (PO^2^) <93%. Reduced level of consciousness was defined as reduced levels of responsiveness to verbal and noxious stimuli from obtundation to coma. Presence of other symptoms, including shivering, loss of smell (anosmia) and taste (ageusia), abdominal pain, nausea, and vomiting were documented by some centers but not systematically inquired.

Comorbidities include Type 2 diabetes mellitus(T2D), cardiovascular disease, acute kidney injury, asthma, currently pregnant, chronic renal failure, cancer, and history of HIV/AIDS. Type 2 diabetes mellitus (T2D) was defined as fasting blood glucose ≥126 mg/dl or use of oral glycemic mediations or insulin. Cardiovascular disease was defined as history of known coronary artery disease by catheterization, history of congestive heart failure according to diagnosis codes, or ejection fraction less than 45%. Acute kidney injury was defined as a drop in glomerular filtration rate (GFR) of 25%. Chronic kidney disease was defined as GFR <60 mL/min/1.73 m^2^. Cancer is defined as active malignancy. Data on the number HIV positive or cancer patients on active therapy is not available.

Intubation was defined as requiring use of a ventilator at any time during hospitalization. Mortality was defined by in-hospital death.

### Study oversight

The study was initiated by The Deputy for Health and Curative affairs at the Iranian Ministry of Health and Medical Education (MOHME) and approved by the institutional review boards of the participating hospitals (http://ethics.research.ac.ir/IndexEn.php). Data collection and analysis was supervised by the Department for Research and Innovation Ministry of Health and Medical Education, Tehran, Iran. The authors have reviewed the data and the manuscript and attest to the accuracy of the data and the adherence to the protocols of the NEJM.org.

### Statistical Analysis

Comparisons between COVID-19 positive to negative patients were made using odds ratios. Cells containing <5 counts were excluded from odds ratio calculations. Comparisons between groups were made using χ^2^ test or Fisher Exact test (when any cell contained ≤10 samples). A 2-tailed p-value <0.05 was considered statistically significant. We conducted data analysis March through April 2020. Data were analyzed using Excel v16.35 (Microsoft) and Stata 16 (StataCorp, LLC).

## RESULTS

### Population Characteristics

A total of 96103 individuals were hospitalized with acute respiratory illness. 18754 (19.5%) tested positive for SARS-CoV-2 by rRT-PC (Table 1; see Supplementary Table 1 for cases by province). The mean age of COVID-19 positive and COVID-19 negative patients were 55.2 and 50.9 years, respectively. There were more men than women among COVID-19 positive patients (61% vs. 39%).

**Table 1:** Total number and percent of COVID-19 positive patients in different age groups

| Age Groups | n | COVID-19 Positive | % |
| --- | --- | --- | --- |
| 0 to 10 years | 2224 | 81 | 3.6% |
| 10 to 20 years | 2094 | 145 | 6.9% |
| 20 to 30 years | 7391 | 836 | 11.3% |
| 30 to 40 years | 14679 | 2553 | 17.4% |
| 40 to 50 years | 15778 | 3274 | 20.8% |
| 50 to 60 years | 17422 | 3878 | 22.3% |
| 60 to 70 years | 16544 | 3664 | 22.1% |
| 70 to 80 years | 11500 | 2543 | 22.1% |
| 80 years and over | 8471 | 1779 | 21.0% |
| Overall | 96103 | 18754 | 19.5% |

Among children under 10 years and 10 to 20 years, 3.6% and 6.8% were COVID-19 positive, respectively (Table 1). The percent positive increased to 11.3% and 17.4% in subjects 20–30 and 30–40 years old, respectively. Above age 40, positive results were present in about 20% of cases (range: 20.8% to 22.3%).

A total of 52666 (54.8%) of the 96103 hospitalized patients were discharged home. 10277 (19.5%) of these 52666 patients were COVID-19 positive (Supplementary Table 1).

### Symptoms and signs

COVID-19 positive patients showed higher incidence of cough, fever, respiratory distress, and myalgia (Table 2, Fig. 1). Whereas, there was a lower incidence of hypoxia and reduced level of consciousness. Directionality of symptoms was similar across all age groups, although differences between COVID-19 positive vs. negative patients were less apparent in the age groups <20 years old (Fig. 1). Additionally, as age increased the frequency of cough, fever, hypoxia decreased while respiratory distress tended to increase (Supplementary Table 2).

**Figure 1:**
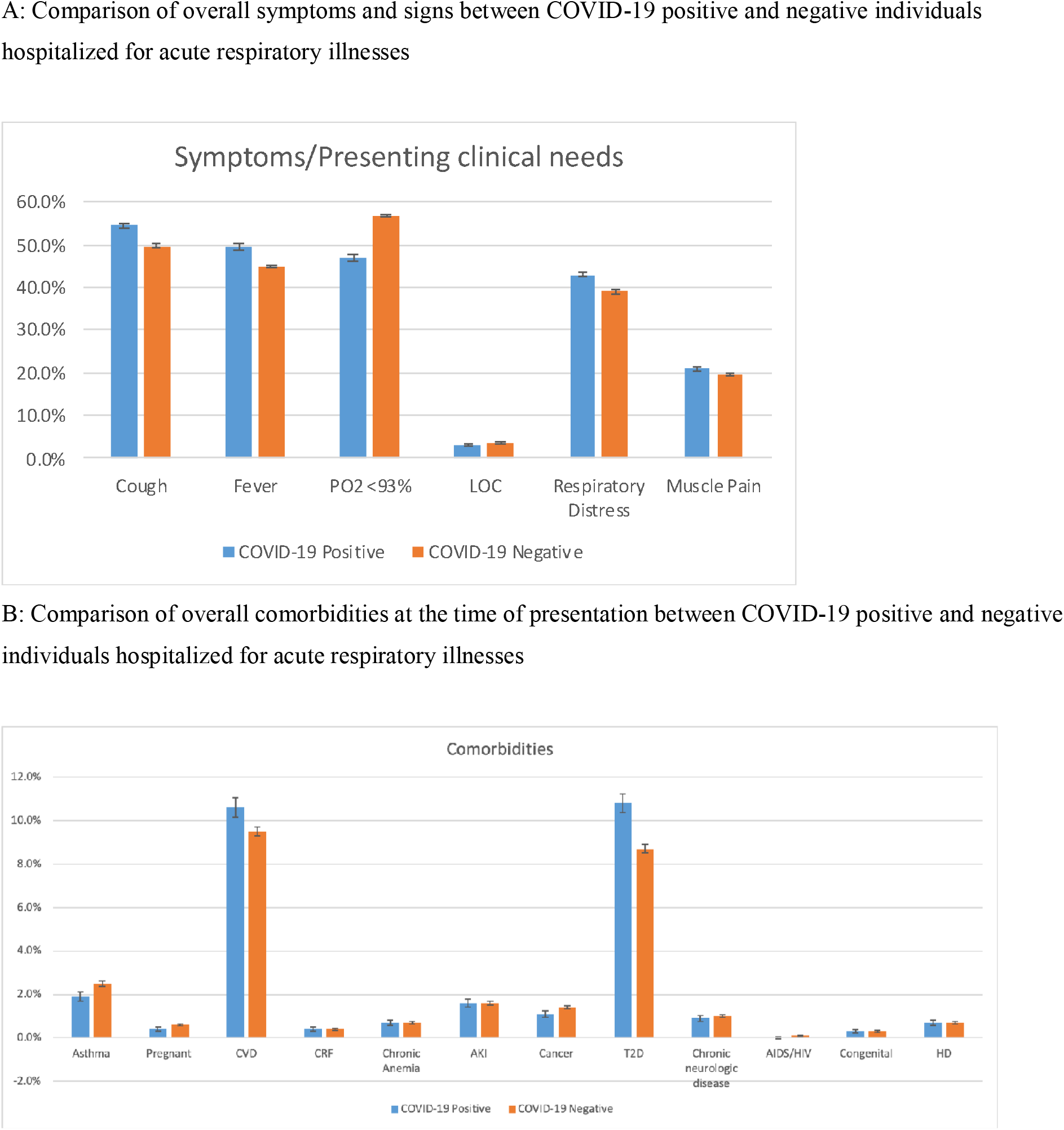
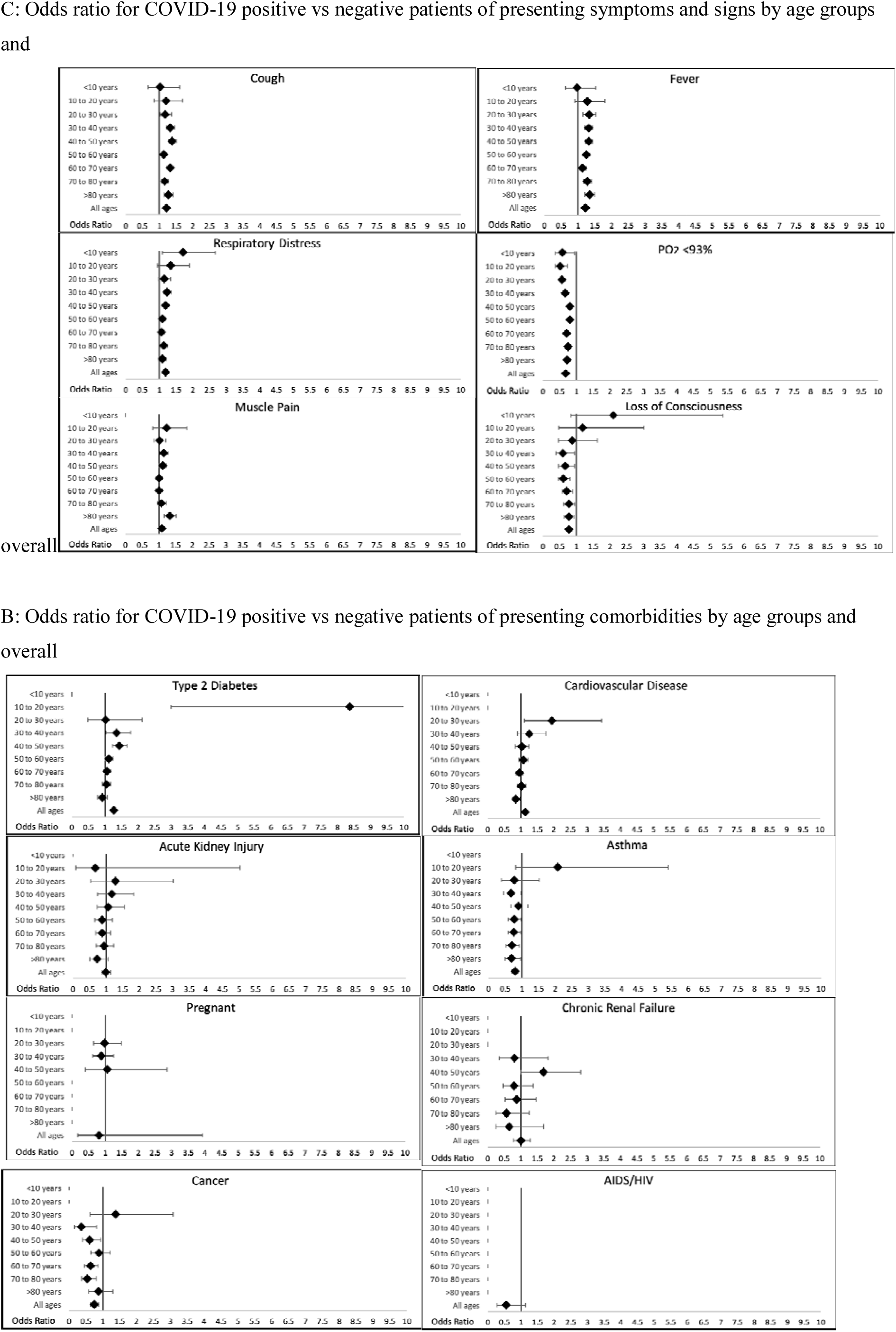
Overall frequency and Odds ratios by age for overall symptoms, signs, and comorbidities between COVID-19 positive and negative individuals hospitalized for acute respiratory illnesses

**Table 2:** Frequency of presenting symptoms and signs, comorbidities, intubation, and death in COVID-19 positive vs negative patients

|  | **Prevalence of**<br>**covariate**<br>**among**<br>**COVID-19**<br>**Positive** | **Prevalence of**<br>**covariate**<br>**among**<br>**COVID-19**<br>**Negative** | **% among those**<br>**reporting**<br>**covariates with**<br>**positive COVID-**<br>**19 result** |
| --- | --- | --- | --- |
| **Symptoms and signs** |  |  |  |
| **Cough** | 54.5% | 49.7% | 21.0% |
| **Fever** | 49.5% | 44.7% | 21.2% |
| **PO2 <93%** | 46.9% | 56.7% | 16.7% |
| **Reduced levels of Consciousness** | 2.8% | 3.6% | 16.0% |
| **Respiratory Distress** | 43.0% | 39.0% | 21.1% |
| **Myalgia** | 20.9% | 19.6% | 20.6% |
| **Comorbidities** |  |  |  |
| **Age (years)** | 55.2 | 50.9 |  |
| **Asthma** | 1.9% | 2.5% | 16.0% |
| **Pregnant** | 0.4% | 0.6% | 13.7% |
| **Cardiovascular Disease** | 10.6% | 9.5% | 21.3% |
| **Chronic kidney disease** | 0.4% | 0.4% | 19.6% |
| **Chronic Anemia** | 0.7% | 0.7% | 19.8% |
| **Acute Kidney Injury** | 1.6% | 1.6% | 19.4% |
| **Cancer** | 1.1% | 1.4% | 15.3% |
| **Type 2 Diabetes Mellitus** | 10.8% | 8.7% | 23.0% |
| **Chronic Neurologic Disease** | 0.9% | 1.0% | 18.1% |
| **AIDS/HIV** | 0.0% | 0.1% | 12.0% |
| **Congenital Diseases** | 0.3% | 0.3% | 21.3% |
| **Hemodialysis** | 0.7% | 0.7% | 18.1% |
| **Ventilator Need** |  |  |  |
| **Intubation** | 7.7% | 5.2% | 26.6% |
| **Outcome** |  |  |  |
| **Death** | 14.6% | 6.3% | 35.9% |

### Comorbidities

T2D and cardiovascular diseases were the two major comorbidities more prevalent among COVID-19 positive compared to COVID-19 negative patients with acute respiratory disease (Table 2. Fig. 1, Supplementary Table 3). There were no significant differences in the rate of HIV/AIDS, chronic kidney disease, chronic anemia, acute kidney injury, hemodialysis treatment and congenital diseases between the two groups. Strikingly there were fewer patients with asthma, cancer, or pregnancy in COVID-19 positive compared to COVID-19 negative patients (Table 2. Fig. 1).

### Intubation/ventilator use

A total of 1450 (7.7%) COVID-19 positive and 3999 (5.2%) COVID-19 negative individuals received ventilatory support (p<0.001) (Supplementary Table 4), which translates to an absolute difference of 26% (Table 2, Fig 2). There were higher number of COVID-19 positive vs. negative patients requiring ventilatory support in all age groups.

**Figure 2:**
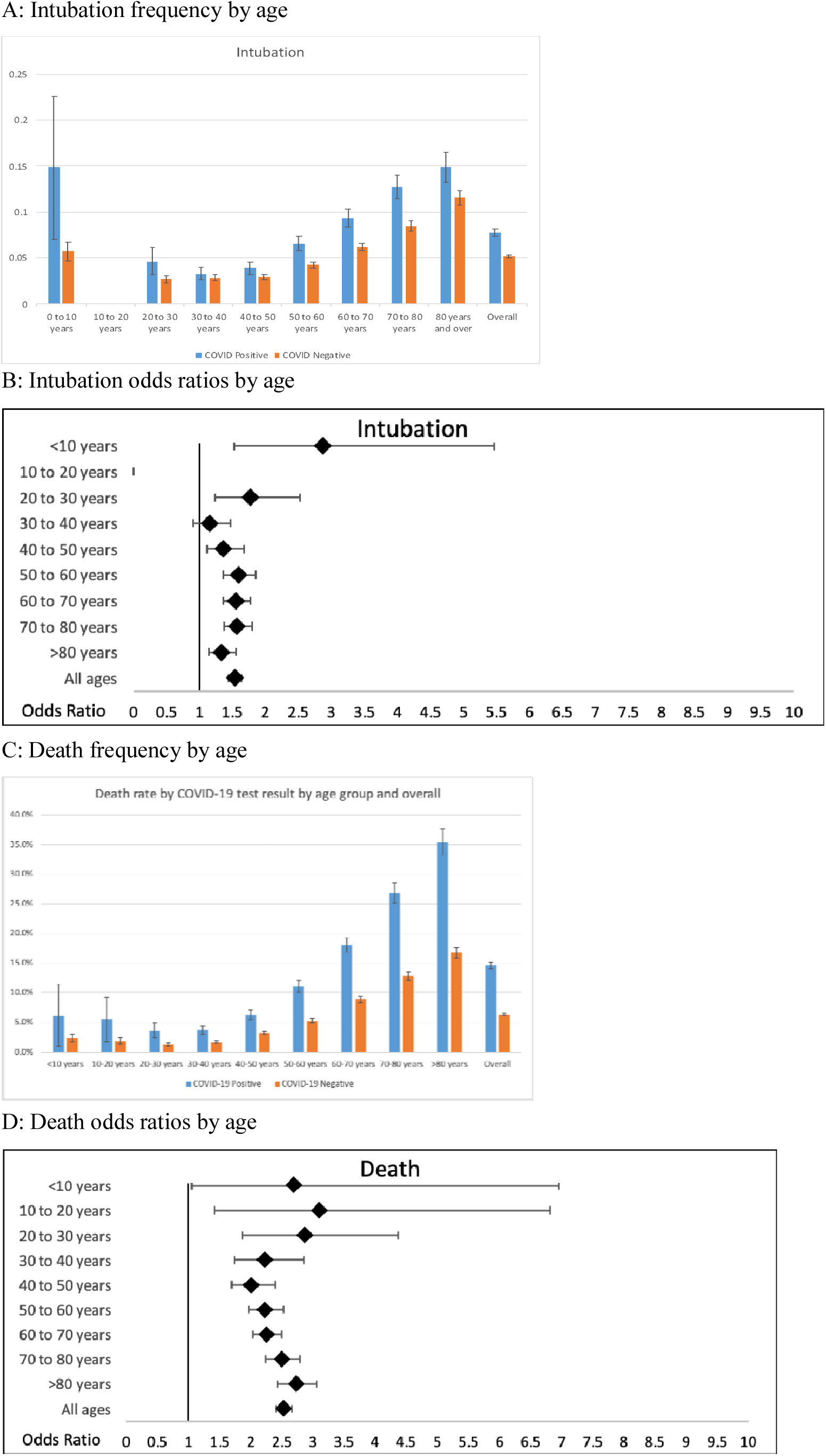
Percentage and odds ratio for intubation and death by COVID-19 results

### Mortality

Frequency of death was higher among COVID-19 positive (n=2740, 14.6%) vs. COVID-19 negative patients (n=4898, 6.3%) (Table 3). The odds ratios for death for COVID-19 positive vs. negative patients was higher across all age groups (Table 3 and Fig. 2).

**Table 3.** Death counts and rate by COVID-19 testing results.

| Age Groups | Death among COVID positive (n) | Death among COVID negative (n) | Death rate among positive cases | Death rate among negative cases | Positive cases among deaths | OR (95% CI) | p-value* | *Note: where there were cells with <10 counts the Fisher Exact test was used, all other tests are $\chi^2$ |
| --- | --- | --- | --- | --- | --- | --- | --- | --- |
| <10 years | 5 | 51 | 6.2% | 2.4% | 8.9% | 2.70 (1.05, 6.95) | 0.051* |  |
| 10 to 20 years | 8 | 36 | 5.5% | 1.8% | 18.2% | 3.10 (1.41, 6.81) | 0.009* |  |
| 20 to 30 years | 30 | 84 | 3.6% | 1.3% | 26.3% | 2.86 (1.88, 4.37) | <0.001 |  |
| 30 to 40 years | 93 | 203 | 3.6% | 1.7% | 31.4% | 2.22 (1.73, 2.85) | <0.001 |  |
| 40 to 50 years | 205 | 402 | 6.3% | 3.2% | 33.8% | 2.01 (1.69, 2.39) | <0.001 |  |
| 50 to 60 years | 427 | 712 | 11.0% | 5.3% | 37.5% | 2.23 (1.97, 2.53) | <0.001 |  |
| 60 to 70 years | 661 | 1146 | 18.0% | 8.9% | 36.6% | 2.25 (2.03, 2.50) | <0.001 |  |
| 70 to 80 years | 681 | 1144 | 26.8% | 12.8% | 37.3% | 2.50 (2.24, 2.78) | <0.001 |  |
| >80 years | 630 | 1120 | 35.4% | 16.7% | 36.0% | 2.73 (2.43, 3.06) | <0.001 |  |
| All ages | 2740 | 4898 | 14.6% | 6.3% | 35.9% | 2.53 (2.41, 2.66) | <0.001 |  |

## DISCUSSION

An urgent collaborative response from multiple Iranian agencies resulted in a rapid symptom screening of a large portion of the country’s population. Among these about 100,000 cases of concern were identified, about 1-in-5 of whom were tested positive for COVID-19 with rRT-PCR. Notably, cases that were COVID-19 positive were more likely to have cough, fever, respiratory distress, and myalgia, but lower level of hypoxia. Positive cases were also more likely to have cardiovascular disease or T2D but less likely to have asthma, cancer, or be pregnant. COVID-19 testing was associated with increased odds for intubation of 1.5 and death of 2.5 times compared to those with negative testing.

The pandemic of COVID-19 has caused devastation in many countries and continues to spread throughout the world. The early diagnosis of the disease in the population is the most efficient way to prevent further spread of this disease and is a necessary step toward its eradication. To this date several types of testing have been generated but none have been perfect. Real-time reverse transcriptase–polymerase chain reaction (rRT-PCR) of nasopharyngeal swabs, targeting the open reading frame *1ab* gene of SARS-CoV-2 have been widely used to confirm the clinical diagnosis[3]. Unfortunately, PCR amplification of virus RNA from nasal swabs have shown low sensitivities in detecting viral infection[1]. While Bronchoalveolar lavage fluid specimens have shown greater than 90% Sensitivity in detecting viral RNA by rRT-PCR rates (14 of 15; 93%), the method is not practical in normal settings and is associated with high risk of disease transmission to the health care providers. At this point rRT-PCR of the virus RNA from nasal swabs remain the most practical way for diagnosis of the disease. Therefore, we examined its utility in disease diagnosis and as a marker of disease severity in more than 90,000 suspicious cases in Iran. By comparing signs and symptoms and complications of COVID-19 test positive patients with those negative for the test we demonstrate here the utility of this test as a marker of disease severity.

Those positive for COVID-19 in our study had differential symptoms compared to COVID-19 negative cases. One would expect that COVID-19 patient would be more likely to present with all viral type symptoms. This was mostly true in our study. However, in opposition to previous reports[4], oxygen desaturation was not more common among COVID-19 positive vs. negative patients. Instead, other clinical findings of fever, cough, and respiratory distress were associated with positive testing. This suggests that hypoxia should not be the only symptom that increases clinician suspicion for COVID-19

Those positive for COVID-19 in our study were more likely to have cardiovascular disease, diabetes, and had higher rates of intubation and death compared to COVID-19 test-negative patients. This may reflect greater virus replication and higher viral load in severe cases as previously shown[5], which expectedly results in easier detection of viral genome and increased disease severity. Interesting, the rates of intubation and death were higher in COVID-19 test-positive vs. test-negative patients in all age groups, including children and adolescents. Earlier studies from China had shown higher rate of disease and its complications among patients older than 65[6]. Our data indicates that COVID-19 has disproportionately affected young individuals in Iran compared to other countries. This may reflect the unique population structure of Iran, which is significantly younger compared to most other countries (World Population Prospects 2019, https://population.un.org/wpp/). In addition, limited access to study drugs and modern therapies due to the economic sanction may have partially accounted for the increased death rate in the younger generations [7]. High prevalence of T2D and metabolic syndrome in Iranians youth may have also increased susceptibility to disease and its complications.[8] Accordingly, there were more patients with T2D in COVID-19 test-positive vs. negative groups. An association between T2D and poor outcome in patients with COVID-19 has been previously reported [9, 10].

Our data also identifies cardiovascular disease as a comorbidity that increases the susceptibility to COVID-19. One intriguing finding of our study is the reduced number of patients that are COVID-19 test positive among those with HIV/AIDS, asthma, cancer, and pregnant women. Based on data from the trials of antiretroviral therapy in Adults Hospitalized patients with severe COVID-19[4] it is unlikely that HIV patients benefited from antiretroviral drugs. It has been hypothesized that a subgroup of patients with severe COVID-19 might suffer from cytokine storm [11]. It is, therefore, more likely that impaired immune system in HIV patients reduces the frequency of cytokine storm. Alternatively, the lower disease rates among higher risk groups is due to a lower threshold for testing in these populations or an increased adherence to social distance among those with known immunocompromising diseases. Information on what percentage of the patients with cancer were being treated with immune suppressive therapy is therefore critical but lacking at this point. In addition, data on lymphocyte count, plasma CRP, IL-6 and TNF was not available when our data were compiled and is being actively collected. Undoubtedly, establishing as to whether cytokine storm underlies disease pathogenesis in severe COVID-19 can have important therapeutic implications.

As with many other countries in the world, lack of experience has undoubtedly led to shortcomings and errors in facing this pandemic. However, considering the structure of the society that promotes high social contacts and many public events that have occurred since the begin of the COVID-19 pandemic mortality in Iran has been significantly lower than anticipated. One possible factor that has limited the number of deaths is the existence of a centralized health care system that has allowed health care authorities to allocate more hospitals and beds and issue uniform guidelines for therapy and use of resources for COVID-19 patients. Accordingly, in contrast to many other countries no shortage of ventilators in Iran has been encountered.

Our study has limitations that confines interpretation. Our data does not include the chest CT-scan results of patients with acute respiratory illness. Access to this data given the probability of false negative PCR results is particularly important. Additionally, the data analyzed in our study were aggregated and hence adjustment for multiple covariates due to lack of access to patient level data could not be carried out. Nevertheless, the higher rate of mortality in patients tested positive vs. those tested negatives suggests that the PCR-based testing has been a relatively suitable method for the screening of high-risk patients.

## CONCLUSION

Our experience suggests that use of rRT-PCR diagnostic method allows better identification of high-risk patients and early utilization of advanced therapeutic measures. Lessons from Iranian experience of COVID-19 should hopefully assist other countries in reducing the fatalities and the economic burden of this pandemic and facilitate preparedness for future pandemics.

## Data Availability

all data presented are available upon request

## ACKNOWLEDGEMENTS

We would like to acknowledge all clinicians, nurses, and healthcare workers in 879 hospitals in Iran for their unlimited devotion to the care of Iranian COVID-19 patients while sacrificing their health and lives. We like to thank Dr. Saeed Namaki, the Minister of health for his leadership and dedication to the care of COVID-19 patients and Drs. Tayeb Ghadimi, and. Abdolkhalegh Keshavarzi, Mr. Reza Mahmoudi Lamouki, and Bagherzadeh for their help with the data preparation, management and analysis.

## Supplementary Appendix

**Supplementary Figure 1:**
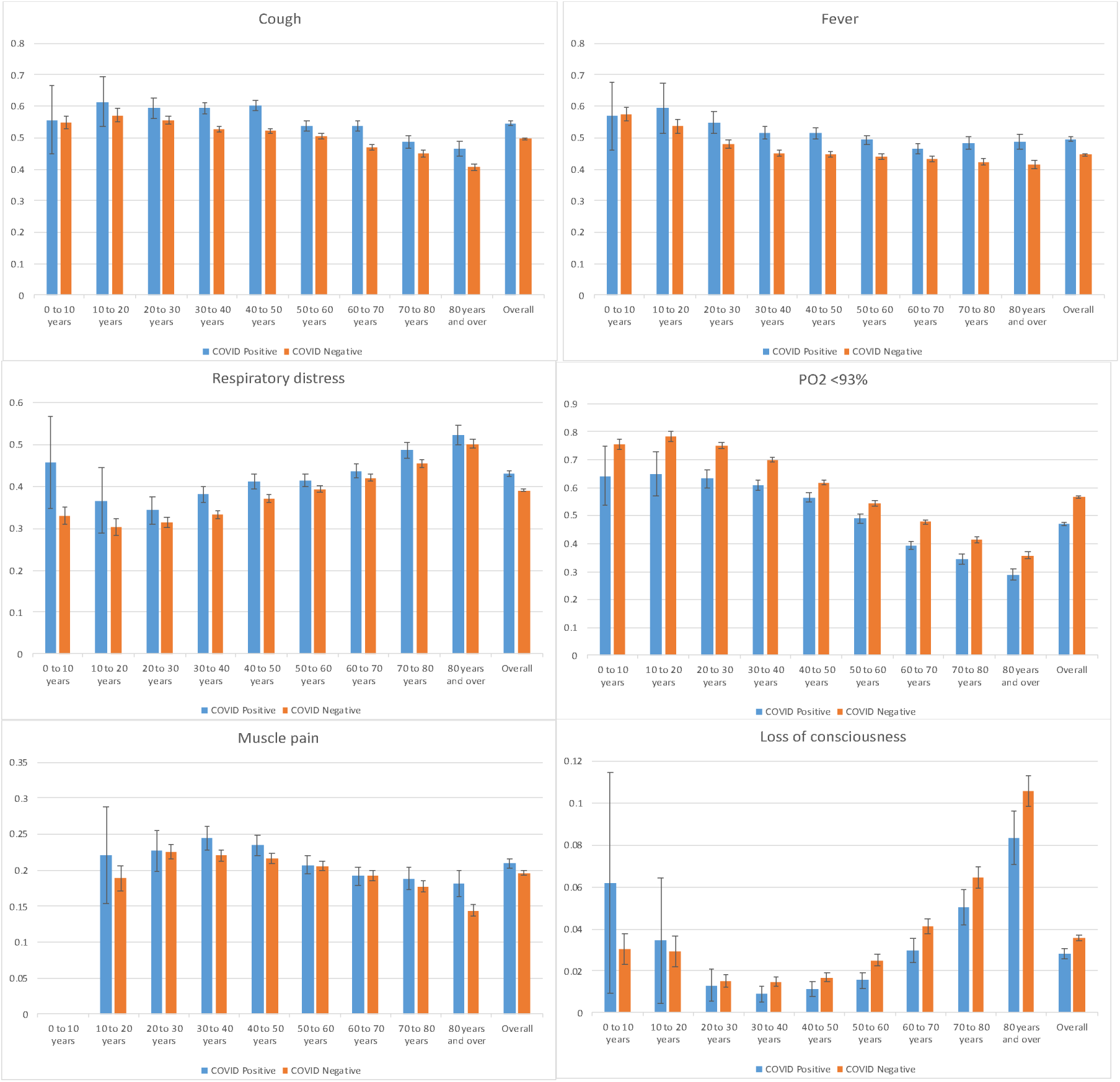
Frequency of symptoms and signs by age groups in COVID-19 Positive vs. Negative patients

**Supplementary Figure 2:**
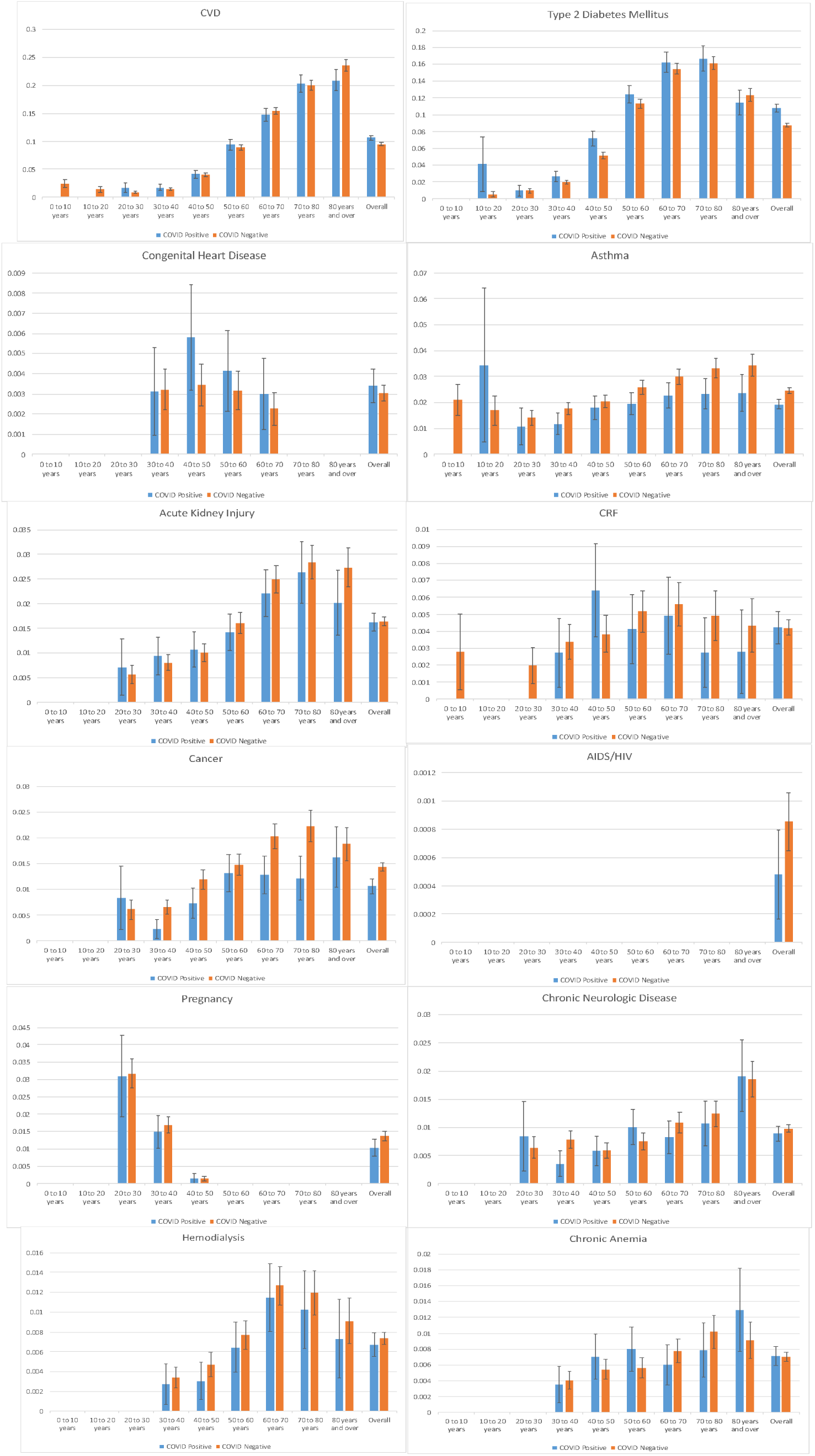
Frequency of comorbidities by age groups in COVID-19 Positive vs. Negative patients

**Supplementary table 1-.** List of total admission for acute respiratory illness during designated month in 31 provinces of Iran and the change in number of admissions between 2 days to one day prior to the completion of the study

| Province | Suspicious cases | Outbreak | Hospitalized | Admitted day-1 | Admitted day-2 | Change |
| --- | --- | --- | --- | --- | --- | --- |
| **Qom Province** | 4873 | 3.7708 | 1969 | 33 | 91 | **-58** |
| **Gilan** | 8019 | 3.1687 | 848 | 86 | 106 | **-20** |
| **Semnan** | 1877 | 2.6724 | 420 | 37 | 60 | **-23** |
| **Mazandaran Province** | 8038 | 2.4479 | 4498 | 128 | 301 | **-173** |
| **Golestan province** | 4264 | 2.2819 | 1344 | 47 | 56 | **-9** |
| **Yazd Province** | 2278 | 2.0008 | 881 | 50 | 110 | **-60** |
| **Tehran Province** | 22549 | 1.6995 | 8957 | 770 | 805 | **-35** |
| **Central Province** | 2254 | 1.5768 | 646 | 42 | 63 | **-21** |
| **Qazvin Province** | 1884 | 1.4791 | 319 | 42 | 48 | **-6** |
| **Isfahan province** | 6606 | 1.29 | 3351 | 156 | 246 | **-90** |
| **Zanjan Province** | 1230 | 1.1632 | 370 | 58 | 60 | **-2** |
| **Lorestan Province** | 1870 | 1.0621 | 303 | 71 | 68 | **3** |
| **Alborz Province** | 2637 | 0.9722 | 1051 | 88 | 110 | **-22** |
| **South Khorasan province** | 696 | 0.9052 | 113 | 15 | 36 | **-21** |
| **Ilam Province** | 517 | 0.8911 | 278 | 22 | 18 | **4** |
| **Bushehr Province** | 959 | 0.8243 | 649 | 15 | 24 | **-9** |
| **Ardebil province** | 987 | 0.7769 | 463 | 30 | 38 | **-8** |
| **Khorasan Razavi Province** | 4584 | 0.7124 | 1218 | 122 | 196 | **-74** |
| **North Khorasan Province** | 612 | 0.7091 | 130 | 15 | 22 | **-7** |
| **Fars province** | 3197 | 0.659 | 668 | 89 | 114 | **-25** |
| **Hamedan province** | 1112 | 0.6324 | 639 | 27 | 55 | **-28** |
| **East Azerbaijan Province** | 2414 | 0.6174 | 1644 | 77 | 87 | **-10** |
| **Khuzestan province** | 2720 | 0.5774 | 1025 | 75 | 105 | **-30** |
| **Hormozgan Province** | 980 | 0.5517 | 76 | 13 | 24 | **-11** |
| **Kurdistan province** | 809 | 0.5047 | 182 | 24 | 34 | **-10** |
| **Kermanshah province** | 905 | 0.4635 | 204 | 18 | 35 | **-17** |
| **West Azerbaijan Province** | 1416 | 0.4337 | 386 | 78 | 64 | **14** |
| **Kerman province** | 1356 | 0.4285 | 281 | 45 | 39 | **6** |
| **Chahar Mahal Bakhtiari Province** | 386 | 0.4073 | 69 | 10 | 13 | **-3** |
| **Sistan and Baluchestan Province** | 1051 | 0.3787 | 181 | 28 | 29 | **-1** |
| **Kohkiluyeh and Boyer Ahmad Province** | 219 | 0.3071 | 100 | 4 | 7 | **-3** |
| **Total** | **93299** | **36.3658** | **33263** | **2315** | **3064** | **-749** |

**Supplement Table 2:** Percent, odds ratios, p-values for symptoms by COVID-19 testing result

| A: Cough |  |  |  |  |
| --- | --- | --- | --- | --- |
| Age Groups | % among positive cases | % among negative cases | Odds Ratio (95% CI) | p-value |
| <10 years | 55.6% | 54.8% | 1.03 (0.66, 1.61) | 0.897 |
| 10 to 20 years | 61.4% | 57.1% | 1.19 (0.84, 1.69) | 0.315 |
| 20 to 30 years | 59.4% | 55.4% | 1.18 (1.02, 1.36) | 0.030 |
| 30 to 40 years | 59.3% | 52.5% | 1.32 (1.21, 1.44) | 0.000 |
| 40 to 50 years | 60.1% | 52.1% | 1.38 (1.28, 1.50) | 0.000 |
| 50 to 60 years | 53.6% | 50.5% | 1.13 (1.06, 1.22) | 0.001 |
| 60 to 70 years | 53.7% | 46.8% | 1.32 (1.23, 1.42) | 0.000 |
| 70 to 80 years | 48.7% | 45.0% | 1.16 (1.06, 1.27) | 0.001 |
| >80 years | 46.4% | 40.6% | 1.27 (1.14, 1.41) | 0.000 |
| All ages | 54.5% | 49.7% | 1.21 (1.18, 1.25) | 0.000 |
| B: Fever |  |  |  |  |
| Age Groups | % among positive cases | % among negative cases | Odds Ratio (95% CI) | p-value |
| <10 years | 56.8% | 57.4% | 0.97 (0.62, 1.52) | 0.907 |
| 10 to 20 years | 59.3% | 53.6% | 1.26 (0.90, 1.78) | 0.181 |
| 20 to 30 years | 54.8% | 47.9% | 1.32 (1.14, 1.52) | 0.000 |
| 30 to 40 years | 51.7% | 45.0% | 1.31 (1.20, 1.42) | 0.000 |
| 40 to 50 years | 51.4% | 44.6% | 1.31 (1.21, 1.42) | 0.000 |
| 50 to 60 years | 49.2% | 43.9% | 1.24 (1.15, 1.33) | 0.000 |
| 60 to 70 years | 46.4% | 43.3% | 1.13 (1.05, 1.22) | 0.001 |
| 70 to 80 years | 48.2% | 42.3% | 1.27 (1.16, 1.38) | 0.000 |
| >80 years | 48.6% | 41.4% | 1.34 (1.20, 1.48) | 0.000 |
| All ages | 49.5% | 44.7% | 1.22 (1.18, 1.26) | 0.000 |
| C: Intubation |  |  |  |  |
| Age Groups | % among positive cases | % among negative cases | Odds Ratio (95% CI) | p-value* |
| <10 years | 14.8% | 5.7% | 2.88 (1.52, 5.46) | 0.001 |
| 10 to 20 years |  |  |  |  |
| 20 to 30 years | 4.7% | 2.7% | 1.77 (1.24, 2.52) | 0.001 |
| 30 to 40 years | 3.3% | 2.9% | 1.15 (0.91, 1.47) | 0.244 |
| 40 to 50 years | 3.9% | 2.9% | 1.36 (1.11, 1.68) | 0.003 |
| 50 to 60 years | 6.5% | 4.2% | 1.59 (1.36, 1.85) | 0.000 |
| 60 to 70 years | 9.3% | 6.2% | 1.55 (1.36, 1.77) | 0.000 |
| 70 to 80 years | 12.7% | 8.5% | 1.57 (1.37, 1.80) | 0.000 |
| >80 years | 14.8% | 11.6% | 1.33 (1.15, 1.55) | 0.000 |
| All ages | 7.7% | 5.2% | 1.54 (1.44, 1.64) | 0.000 |
| D: PO2 <93% |  |  |  |  |
| Age Groups | % among positive cases | % among negative cases | Odds Ratio (95% CI) | p-value |
| <10 years | 64.2% | 75.3% | 0.59 (0.37, 0.94) | 0.024 |
| 10 to 20 years | 64.8% | 78.1% | 0.52 (0.36, 0.74) | 0.000 |
| 20 to 30 years | 63.2% | 75.0% | 0.57 (0.49, 0.67) | 0.000 |
| 30 to 40 years | 60.9% | 69.9% | 0.67 (0.61, 0.73) | 0.000 |
| 40 to 50 years | 56.4% | 61.7% | 0.80 (0.74, 0.87) | 0.000 |
| 50 to 60 years | 48.9% | 54.3% | 0.80 (0.75, 0.86) | 0.000 |
| 60 to 70 years | 39.3% | 47.7% | 0.71 (0.66, 0.76) | 0.000 |
| 70 to 80 years | 34.5% | 41.2% | 0.75 (0.69, 0.82) | 0.000 |
| >80 years | 28.9% | 35.9% | 0.73 (0.65, 0.81) | 0.000 |
| All ages | 46.9% | 56.7% | 0.68 (0.66, 0.70) | 0.000 |
| E: Reduced Levels of consciousness |  |  |  |  |
| Age Groups | % among positive cases | % among negative cases | Odds Ratio (95% CI) | p-value* |
| <10 years | 6.2% | 3.0% | 2.10 (0.82, 5.37) | 0.109 |
| 10 to 20 years | 3.4% | 2.9% | 1.19 (0.47, 3.01) | 0.615 |
| 20 to 30 years | 1.3% | 1.5% | 0.87 (0.46, 1.63) | 0.659 |
| 30 to 40 years | 0.9% | 1.5% | 0.60 (0.39, 0.93) | 0.022 |
| 40 to 50 years | 1.1% | 1.7% | 0.66 (0.47, 0.94) | 0.021 |
| 50 to 60 years | 1.5% | 2.5% | 0.61 (0.46, 0.81) | 0.000 |
| 60 to 70 years | 3.0% | 4.1% | 0.71 (0.58, 0.88) | 0.001 |
| 70 to 80 years | 5.0% | 6.5% | 0.77 (0.63, 0.94) | 0.008 |
| >80 years | 8.3% | 10.5% | 0.77 (0.64, 0.93) | 0.006 |
| All ages | 2.8% | 3.6% | 0.78 (0.71, 0.86) | 0.000 |
| F: Respiratory distress |  |  |  |  |
| Age Groups | % among positive cases | % among negative cases | Odds Ratio (95% CI) | p-value |
| <10 years | 45.7% | 32.9% | 1.71 (1.10, 2.67) | 0.017 |
| 10 to 20 years | 36.6% | 30.2% | 1.33 (0.94, 1.90) | 0.108 |
| 20 to 30 years | 34.3% | 31.4% | 1.14 (0.98, 1.33) | 0.086 |
| 30 to 40 years | 38.0% | 33.3% | 1.23 (1.13, 1.34) | 0.000 |
| 40 to 50 years | 41.1% | 37.1% | 1.19 (1.10, 1.28) | 0.000 |
| 50 to 60 years | 41.4% | 39.3% | 1.09 (1.01, 1.17) | 0.018 |
| 60 to 70 years | 43.6% | 42.0% | 1.07 (0.99, 1.15) | 0.081 |
| 70 to 80 years | 48.6% | 45.4% | 1.14 (1.04, 1.24) | 0.004 |
| >80 years | 52.2% | 50.1% | 1.08 (0.98, 1.20) | 0.131 |
| All ages | 43.0% | 39.0% | 1.18 (1.14, 1.22) | 0.000 |
| G: Myalgia |  |  |  |  |
| Age Groups | % among positive cases | % among negative cases | Odds Ratio (95% CI) | p-value* |
| <10 years |  |  |  |  |
| 10 to 20 years | 22.1% | 18.9% | 1.22 (0.81, 1.83) | 0.346 |
| 20 to 30 years | 22.7% | 22.5% | 1.01 (0.85, 1.20) | 0.915 |
| 30 to 40 years | 24.4% | 22.0% | 1.14 (1.03, 1.26) | 0.009 |
| 40 to 50 years | 23.4% | 21.6% | 1.11 (1.01, 1.21) | 0.027 |
| 50 to 60 years | 20.7% | 20.6% | 1.01 (0.92, 1.10) | 0.880 |
| 60 to 70 years | 19.2% | 19.2% | 1.00 (0.91, 1.10) | 0.985 |
| 70 to 80 years | 18.8% | 17.7% | 1.08 (0.96, 1.21) | 0.195 |
| >80 years | 18.2% | 14.4% | 1.32 (1.15, 1.51) | 0.000 |
| All ages | 20.9% | 19.6% | 1.08 (1.04, 1.13) | 0.000 |

\* Note: where there were cells with <10 counts the Fisher Exact test was used, all other tests are $\chi^2$ .

**Supplement Table 3:** Percent, odds ratio, p-values for comorbidities by COVID-19 testing results

| A: Asthma |  |  |  |  |
| --- | --- | --- | --- | --- |
| Age Groups | % among positive cases | % among negative cases | Odds Ratio (95% CI) | p-value* |
| <10 years |  |  |  |  |
| 10 to 20 years | 3.4% | 1.7% | 2.07 (0.80, 5.39) | 0.180 |
| 20 to 30 years | 1.1% | 1.4% | 0.76 (0.38, 1.52) | 0.530 |
| 30 to 40 years | 1.2% | 1.8% | 0.66 (0.45, 0.97) | 0.034 |
| 40 to 50 years | 1.8% | 2.0% | 0.88 (0.66, 1.17) | 0.372 |
| 50 to 60 years | 2.0% | 2.6% | 0.76 (0.59, 0.97) | 0.028 |
| 60 to 70 years | 2.3% | 3.0% | 0.75 (0.59, 0.95) | 0.017 |
| 70 to 80 years | 2.3% | 3.3% | 0.69 (0.52, 0.92) | 0.010 |
| >80 years | 2.4% | 3.4% | 0.68 (0.49, 0.95) | 0.022 |
| All ages | 1.9% | 2.5% | 0.78 (0.70, 0.88) | 0.000 |
| B: Pregnant |  |  |  |  |
| Age Groups | % among positive cases | % among negative cases | Odds Ratio (95% CI) | alue* |
| <10 years |  |  |  |  |
| 10 to 20 years |  |  |  |  |
| 20 to 30 years | 3.1% | 3.2% | 0.98 (0.65, 1.48) | 0.917 |
| 30 to 40 years | 1.5% | 1.7% | 0.88 (0.62, 1.25) | 0.484 |
| 40 to 50 years | 0.2% | 0.1% | 1.06 (0.39, 2.86) | 0.802 |
| 50 to 60 years |  |  |  |  |
| 60 to 70 years |  |  |  |  |
| 70 to 80 years |  |  |  |  |
| >80 years |  |  |  |  |
| All ages | 0.4% | 0.6% | 0.81 (0.17, 3.92) | 0.001 |
| C: Cardiovascular Disease |  |  |  |  |
| Age Groups | % among positive cases | % among negative cases | Odds Ratio (95% CI) | p-value* |
| <10 years |  |  |  |  |
| 10 to 20 years |  |  |  |  |
| 20 to 30 years | 1.8% | 0.9% | 1.94 (1.10, 3.43) | 0.020 |
| 30 to 40 years | 1.8% | 1.4% | 1.25 (0.90, 1.74) | 0.176 |
| 40 to 50 years | 4.2% | 4.1% | 1.02 (0.84, 1.24) | 0.814 |
| 50 to 60 years | 9.4% | 8.9% | 1.07 (0.94, 1.21) | 0.302 |
| 60 to 70 years | 14.8% | 15.4% | 0.95 (0.86, 1.06) | 0.379 |
| 70 to 80 years | 20.3% | 20.0% | 1.02 (0.91, 1.13) | 0.776 |
| >80 years | 20.9% | 23.5% | 0.86 (0.76, 0.97) | 0.019 |
| All ages | 10.6% | 9.5% | 1.13 (1.07, 1.19) | 0.000 |
| D: Chronic Renal Failure |  |  |  |  |
| Age Groups | % among positive cases | % among negative cases | Odds Ratio (95% CI) | p-value* |
| <10 years |  |  |  |  |
| 10 to 20 years |  |  |  |  |
| 20 to 30 years |  |  |  |  |
| 30 to 40 years | 0.3% | 0.3% | 0.81 (0.36, 1.81) | 0.706 |
| 40 to 50 years | 0.6% | 0.4% | 1.68 (1.00, 2.80) | 0.047 |
| 50 to 60 years | 0.4% | 0.5% | 0.80 (0.46, 1.37) | 0.414 |
| 60 to 70 years | 0.5% | 0.6% | 0.88 (0.52, 1.47) | 0.623 |
| 70 to 80 years | 0.3% | 0.5% | 0.56 (0.25, 1.24) | 0.177 |
| >80 years | 0.3% | 0.4% | 0.65 (0.25, 1.68) | 0.526 |
| All ages | 0.4% | 0.4% | 1.00 (0.78, 1.28) | 0.984 |
| E: Chronic anemia |  |  |  |  |
| Age Groups | % among positive cases | % among negative cases | Odds Ratio (95% CI) | p-value* |
| <10 years |  |  |  |  |
| 10 to 20 years |  |  |  |  |
| 20 to 30 years |  |  |  |  |
| 30 to 40 years | 0.4% | 0.4% | 0.87 (0.43, 1.78) | 0.862 |
| 40 to 50 years | 0.7% | 0.5% | 1.29 (0.81, 2.08) | 0.286 |
| 50 to 60 years | 0.8% | 0.6% | 1.43 (0.94, 2.17) | 0.094 |
| 60 to 70 years | 0.6% | 0.8% | 0.77 (0.49, 1.23) | 0.272 |
| 70 to 80 years | 0.8% | 1.0% | 0.77 (0.48, 1.26) | 0.296 |
| >80 years | 1.3% | 0.9% | 1.42 (0.88, 2.31) | 0.149 |
| All ages | 0.7% | 0.7% | 1.02 (0.84, 1.23) | 0.869 |
| F: Acute Kidney Injury |  |  |  |  |
| Age Groups | % among positive cases | % among negative cases | Odds Ratio (95% CI) | p-value* |
| <10 years |  |  |  |  |
| 10 to 20 years | 0.7% | 1.0% | 0.67 (0.09, 5.03) | 1.000 |
| 20 to 30 years | 0.7% | 0.6% | 1.27 (0.54, 3.02) | 0.626 |
| 30 to 40 years | 0.9% | 0.8% | 1.16 (0.74, 1.82) | 0.505 |
| 40 to 50 years | 1.1% | 1.0% | 1.06 (0.73, 1.55) | 0.756 |
| 50 to 60 years | 1.4% | 1.6% | 0.88 (0.65, 1.18) | 0.398 |
| 60 to 70 years | 2.2% | 2.5% | 0.88 (0.69, 1.13) | 0.329 |
| 70 to 80 years | 2.6% | 2.8% | 0.93 (0.71, 1.22) | 0.587 |
| >80 years | 2.0% | 2.7% | 0.73 (0.51, 1.05) | 0.093 |
| All ages | 1.6% | 1.6% | 0.99 (0.88, 1.13) | 0.931 |
| G: Cancer |  |  |  |  |
| Age Groups | % among positive cases | % among negative cases | Odds Ratio (95% CI) | p-value* |
| <10 years |  |  |  |  |
| 10 to 20 years |  |  |  |  |
| 20 to 30 years | 0.8% | 0.6% | 1.37 (0.61, 3.08) | 0.480 |
| 30 to 40 years | 0.2% | 0.7% | 0.35 (0.15, 0.81) | 0.009 |
| 40 to 50 years | 0.7% | 1.2% | 0.61 (0.39, 0.94) | 0.023 |
| 50 to 60 years | 1.3% | 1.5% | 0.88 (0.65, 1.21) | 0.437 |
| 60 to 70 years | 1.3% | 2.0% | 0.63 (0.46, 0.86) | 0.003 |
| 70 to 80 years | 1.2% | 2.2% | 0.54 (0.37, 0.79) | 0.001 |
| >80 years | 1.6% | 1.9% | 0.86 (0.57, 1.30) | 0.480 |
| All ages | 1.1% | 1.4% | 0.74 (0.64, 0.86) | 0.000 |
| H: Type 2 diabetes mellitus |  |  |  |  |
| Age Groups | % among positive cases | % among negative cases | Odds Ratio (95% CI) | p-value* |
| <10 years |  |  |  |  |
| 10 to 20 years | 4.1% | 0.5% | 8.37 (3.00, 23.37) | <0.001 |
| 20 to 30 years | 1.0% | 0.9% | 1.01 (0.48, 2.12) | 1.000 |
| 30 to 40 years | 2.7% | 2.0% | 1.34 (1.02, 1.76) | 0.033 |
| 40 to 50 years | 7.2% | 5.1% | 1.43 (1.22, 1.67) | 0.000 |
| 50 to 60 years | 12.4% | 11.3% | 1.11 (1.00, 1.24) | 0.055 |
| 60 to 70 years | 16.2% | 15.4% | 1.06 (0.96, 1.17) | 0.253 |
| 70 to 80 years | 16.7% | 16.2% | 1.04 (0.92, 1.17) | 0.541 |
| >80 years | 11.4% | 12.3% | 0.91 (0.78, 1.08) | 0.285 |
| All ages | 10.8% | 8.7% | 1.26 (1.20, 1.33) | 0.000 |
| I: Chronic neurologic disease |  |  |  |  |
| Age Groups | % among positive cases | % among negative cases | Odds Ratio (95% CI) | p-value* |
| <10 years | 1.2% | 1.9% | 0.66 (0.09, 4.84) | 1.000 |
| 10 to 20 years | 0.7% | 1.5% | 0.46 (0.06, 3.40) | 0.718 |
| 20 to 30 years | 0.8% | 0.6% | 1.31 (0.59, 2.92) | 0.495 |
| 30 to 40 years | 0.4% | 0.8% | 0.45 (0.23, 0.89) | 0.019 |
| 40 to 50 years | 0.6% | 0.6% | 0.98 (0.59, 1.63) | 0.939 |
| 50 to 60 years | 1.0% | 0.8% | 1.34 (0.92, 1.94) | 0.122 |
| 60 to 70 years | 0.8% | 1.1% | 0.76 (0.51, 1.12) | 0.167 |
| 70 to 80 years | 1.1% | 1.2% | 0.86 (0.56, 1.31) | 0.468 |
| >80 years | 1.9% | 1.9% | 1.03 (0.70, 1.51) | 0.872 |
| All ages | 0.9% | 1.0% | 0.91 (0.77, 1.08) | 0.274 |
| J: AIDS or HIV |  |  |  |  |
| Age Groups | % among positive cases | % among negative cases | Odds Ratio (95% CI) | p-value* |
| <10 years |  |  |  |  |
| 10 to 20 years |  |  |  |  |
| 20 to 30 years |  |  |  |  |
| 30 to 40 years |  |  |  |  |
| 40 to 50 years |  |  |  |  |
| 50 to 60 years |  |  |  |  |
| 60 to 70 years |  |  |  |  |
| 70 to 80 years |  |  |  |  |
| >80 years |  |  |  |  |
| All ages | 0.0% | 0.1% | 0.56 (0.28, 1.13) | 0.100 |
| K: Congenital heart disease |  |  |  |  |
| Age Groups | % among positive cases | % among negative cases | Odds Ratio (95% CI) | p-value* |
| <10 years |  |  |  |  |
| 10 to 20 years |  |  |  |  |
| 20 to 30 years |  |  |  |  |
| 30 to 40 years | 0.3% | 0.3% | 0.97 (0.45, 2.09) | 1.000 |
| 40 to 50 years | 0.6% | 0.3% | 1.69 (0.98, 2.91) | 0.054 |
| 50 to 60 years | 0.4% | 0.3% | 1.30 (0.73, 2.31) | 0.369 |
| 60 to 70 years | 0.3% | 0.2% | 1.33 (0.67, 2.67) | 0.414 |
| 70 to 80 years | 0.2% | 0.2% | 1.01 (0.33, 3.06) | 1.000 |
| >80 years | 0.1% | 0.2% | 0.27 (0.04, 2.04) | 0.220 |
| All ages | 0.3% | 0.3% | 1.12 (0.85, 1.48) | 0.426 |
| L: Hemodialysis |  |  |  |  |
| Age Groups | % among positive cases | % among negative cases | Odds Ratio (95% CI) | p-value* |
| <10 years | 1.2% | 0.1% | 8.92 (0.92, 86.67) | 0.138 |
| 10 to 20 years | 0.7% | 0.5% | 1.50 (0.19, 11.90) | 0.513 |
| 20 to 30 years | 0.1% | 0.4% | 0.34 (0.05, 2.52) | 0.512 |
| 30 to 40 years | 0.3% | 0.3% | 0.81 (0.36, 1.81) | 0.706 |
| 40 to 50 years | 0.3% | 0.5% | 0.65 (0.33, 1.26) | 0.235 |
| 50 to 60 years | 0.6% | 0.8% | 0.84 (0.54, 1.30) | 0.430 |
| 60 to 70 years | 1.1% | 1.3% | 0.90 (0.64, 1.27) | 0.565 |
| 70 to 80 years | 1.0% | 1.2% | 0.85 (0.56, 1.31) | 0.474 |
| >80 years | 0.7% | 0.9% | 0.80 (0.44, 1.46) | 0.466 |
| All ages | 0.7% | 0.7% | 0.91 (0.75, 1.11) | 0.346 |

\* Note: where there were cells with <10 counts the Fisher Exact test was used, all other tests are $\chi^2$ .

**Supplement Table 4:**
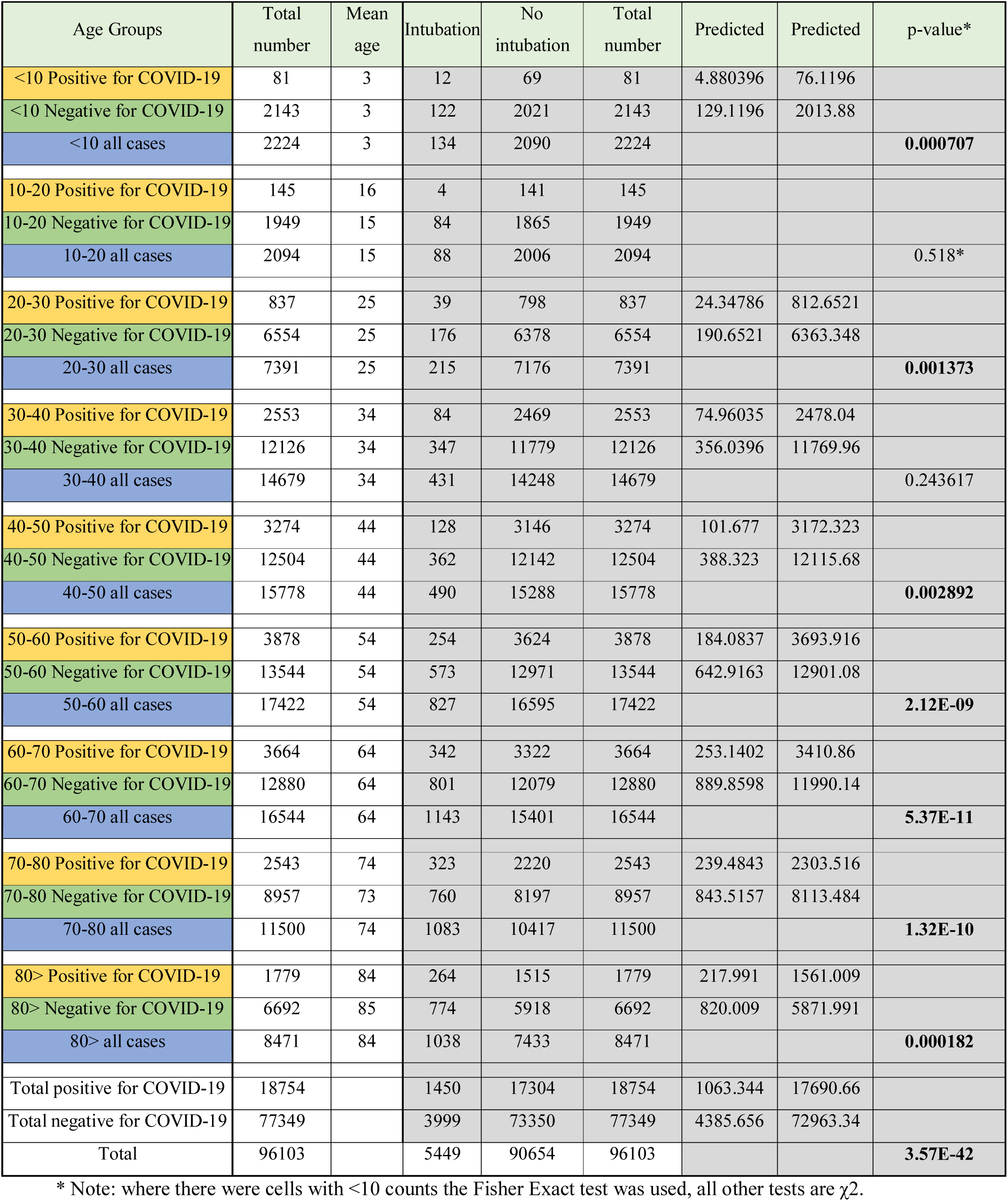
Intubation by age group, including calculations for χ^2^ testing.

**Supplement Table 5:**
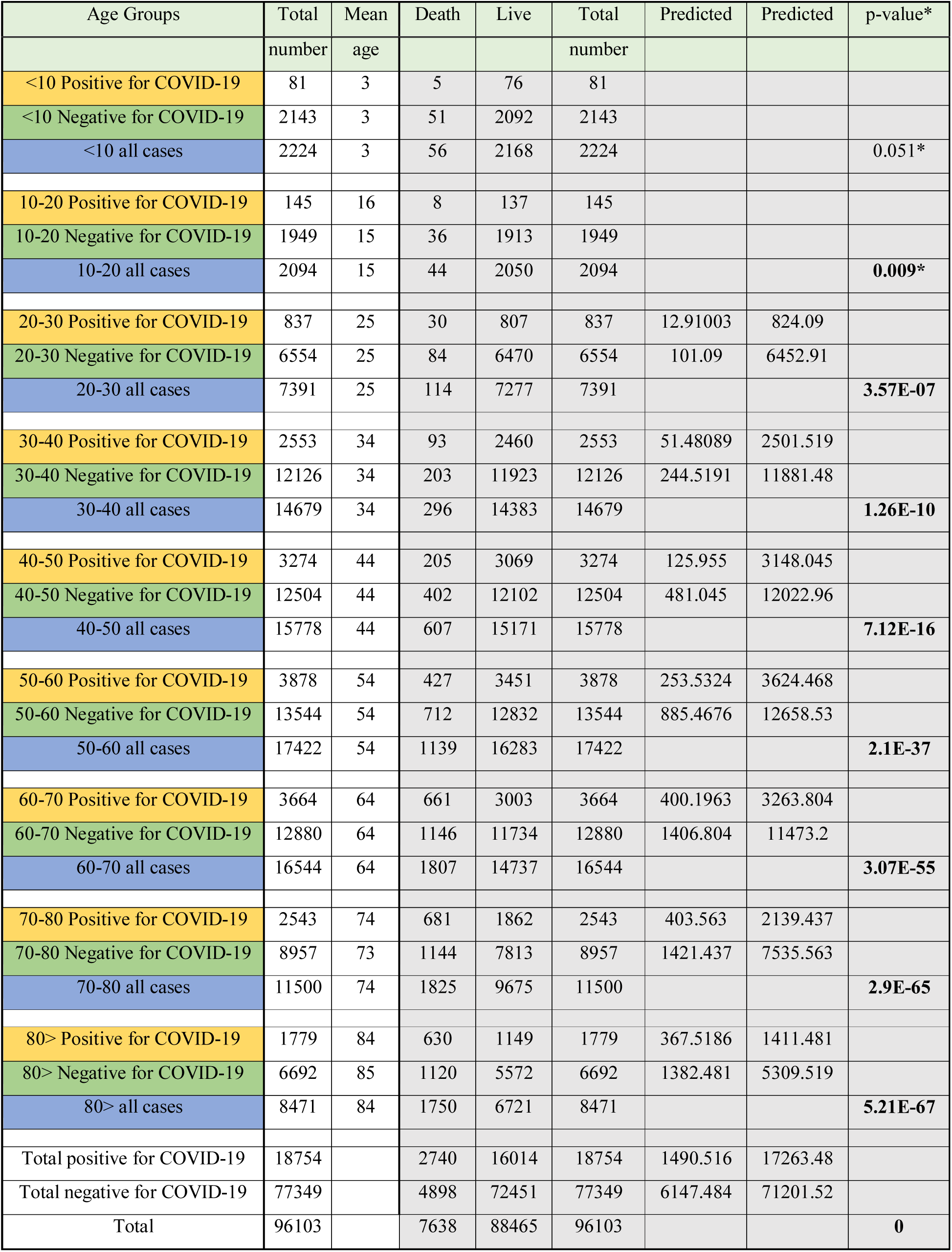
Death by age group, including calculations for χ^2^ testing.

| Age Groups | Total | Mean | Death | Live | Total | Predicted | Predicted | p-value* |
| --- | --- | --- | --- | --- | --- | --- | --- | --- |

|  | number | age |  |  | number |  |  |  |
| --- | --- | --- | --- | --- | --- | --- | --- | --- |
| <10 Positive for COVID-19 | 81 | 3 | 5 | 76 | 81 |  |  |  |
| <10 Negative for COVID-19 | 2143 | 3 | 51 | 2092 | 2143 |  |  |  |
| <10 all cases | 2224 | 3 | 56 | 2168 | 2224 |  |  | 0.051* |
| 10-20 Positive for COVID-19 | 145 | 16 | 8 | 137 | 145 |  |  |  |
| 10-20 Negative for COVID-19 | 1949 | 15 | 36 | 1913 | 1949 |  |  |  |
| 10-20 all cases | 2094 | 15 | 44 | 2050 | 2094 |  |  | 0.009* |
| 20-30 Positive for COVID-19 | 837 | 25 | 30 | 807 | 837 | 12.91003 | 824.09 |  |
| 20-30 Negative for COVID-19 | 6554 | 25 | 84 | 6470 | 6554 | 101.09 | 6452.91 |  |
| 20-30 all cases | 7391 | 25 | 114 | 7277 | 7391 |  |  | 3.57E-07 |
| 30-40 Positive for COVID-19 | 2553 | 34 | 93 | 2460 | 2553 | 51.48089 | 2501.519 |  |
| 30-40 Negative for COVID-19 | 12126 | 34 | 203 | 11923 | 12126 | 244.5191 | 11881.48 |  |
| 30-40 all cases | 14679 | 34 | 296 | 14383 | 14679 |  |  | 1.26E-10 |
| 40-50 Positive for COVID-19 | 3274 | 44 | 205 | 3069 | 3274 | 125.955 | 3148.045 |  |
| 40-50 Negative for COVID-19 | 12504 | 44 | 402 | 12102 | 12504 | 481.045 | 12022.96 |  |
| 40-50 all cases | 15778 | 44 | 607 | 15171 | 15778 |  |  | 7.12E-16 |
| 50-60 Positive for COVID-19 | 3878 | 54 | 427 | 3451 | 3878 | 253.5324 | 3624.468 |  |
| 50-60 Negative for COVID-19 | 13544 | 54 | 712 | 12832 | 13544 | 885.4676 | 12658.53 |  |
| 50-60 all cases | 17422 | 54 | 1139 | 16283 | 17422 |  |  | 2.1E-37 |
| 60-70 Positive for COVID-19 | 3664 | 64 | 661 | 3003 | 3664 | 400.1963 | 3263.804 |  |
| 60-70 Negative for COVID-19 | 12880 | 64 | 1146 | 11734 | 12880 | 1406.804 | 11473.2 |  |
| 60-70 all cases | 16544 | 64 | 1807 | 14737 | 16544 |  |  | 3.07E-55 |
| 70-80 Positive for COVID-19 | 2543 | 74 | 681 | 1862 | 2543 | 403.563 | 2139.437 |  |
| 70-80 Negative for COVID-19 | 8957 | 73 | 1144 | 7813 | 8957 | 1421.437 | 7535.563 |  |
| 70-80 all cases | 11500 | 74 | 1825 | 9675 | 11500 |  |  | 2.9E-65 |
| 80> Positive for COVID-19 | 1779 | 84 | 630 | 1149 | 1779 | 367.5186 | 1411.481 |  |
| 80> Negative for COVID-19 | 6692 | 85 | 1120 | 5572 | 6692 | 1382.481 | 5309.519 |  |
| 80> all cases | 8471 | 84 | 1750 | 6721 | 8471 |  |  | 5.21E-67 |
| Total positive for COVID-19 | 18754 |  | 2740 | 16014 | 18754 | 1490.516 | 17263.48 |  |
| Total negative for COVID-19 | 77349 |  | 4898 | 72451 | 77349 | 6147.484 | 71201.52 |  |
| Total | 96103 |  | 7638 | 88465 | 96103 |  |  | 0 |

**Supplement Table 6:**
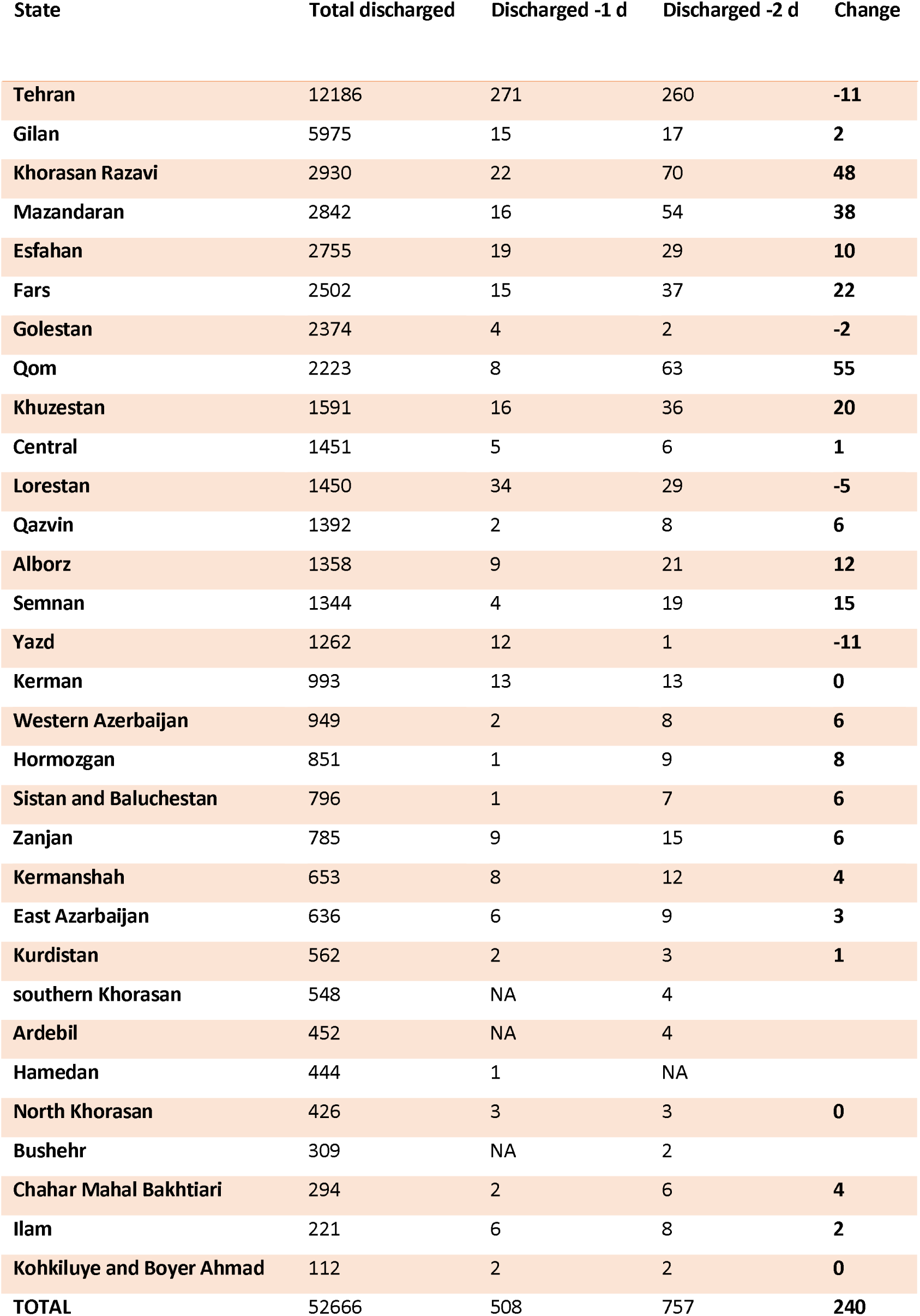
Trend in hospital discharge in 31 provinces; 1 and 2 days prior to completion of the study

## Notes

### Competing Interest Statement

The authors have declared no competing interest.

### Funding Statement

This manuscript was in part funded by a NIH/NHLBI award RHL135767A (NHLBI Outstanding Investigator Award)to A.M.

